# Spatial patterns and determinants of Anemia in women of reproductive age in Zambia (2018–2024): A multilevel ordinal regression approach

**DOI:** 10.64898/2026.03.30.26349744

**Authors:** James Muchinga, Given Moonga, Nawa Mukumbuta, Patrick Musonda

**Affiliations:** Department of Epidemiology and Biostatistics, University of Zambia, Lusaka, Zambia; Department of Natural Sciences, Institute of Basic and Biomedical Sciences, Levy Mwanawasa Medical University, Lusaka, Zambia; Department of Epidemiology and Biostatistics, School of Public Health and Environmental Sciences, Levy Mwanawasa Medical University, Lusaka, Zambia

## Abstract

**Background:** Anemia is a condition characterized by nutritional deficiencies and blood disorders, predominantly affecting children aged 6 to 59 months and women of reproductive age, especially in low and middle-income countries. In Zambia, anemia is a public health problem. This study aims to assess the spatial patterns and determine factors associated with anemia severity in Zambia over six years (2018 to 2024).

**Method:** The study included a total of 19,362 WRA from the two waves of the ZDHS, 2018 and 2024. The ZDHS is a periodic national survey that uses multistage sampling. We adopted an analytical cross-sectional design, and the three-level multivariable ordinal logistic regression model was used to identify variables (individual, household, and community level) associated with anemia severity. Global Moran’s I, Local Moran’s I, and Getis-Ord Gi* statistics were used to determine the hotspots and spatial patterns, while spatial scan statistics were used to detect primary and secondary clusters and their distribution over the two cycles.

**Results:** The prevalence of anemia among women of reproductive age in Zambia was 31.0% (n=3,946) and 30.4% (n=2,015) in 2018 and 2024, respectively. The factors associated with higher odds of anemia severity were HIV status (HIV-positive: AOR=2.63, 95% CI:2.25,3.09), pregnancy (AOR=1.96, 95% CI:1.67,2.31), and rural residency (AOR=1.21, 95% CI:1.08,1.35). While being in a union was protective compared to never being in a union (AOR=0.66, 95% CI:0.57,0.77), not having financial barriers for medical assistance was equally protective. Spatial analysis showed geographic disparities and a non-random distribution of anemia (Global Moran’s I, 2018: I=0.147, p<0.001; 2024: I=0.130, p<0.001). the Hotspot analysis depicted an expansion of high-risk areas Western in 2018 to the North-Western and Luapula in 2024. Spatial scan analysis identified the south-west region (Western, Southern and North-Western) as the significant primary cluster of anemia consistently for both waves.

**Conclusion:** Anemia among women of reproductive age remains a persistent and significant moderate public health problem among WRA in Zambia. Anemia severity is influenced by complex interactions at the individual, household, and community levels. Subtle changes in the effects of predictors were observed over the study period. These finding suggest the need for subnational (provincial and district) targeted interventions, especially in the persistent hotspot regions such as Western and emergine high-risk areas like Luapula, where integrated malaria control, nutritional supplementation, and improved healthcare access should be a priority.

## Introduction

Anemia is defined as a condition in which an individual has insufficient hemoglobin levels, leading to inadequate oxygen supply to the body tissues [1]. This public health issue is particularly pressing in low-and middle-income countries (LMICs), where it is exacerbated by nutritional deficiencies, infectious diseases, and limited healthcare access [2]. Approximately 1.76 billion (about a quarter) of the global population are affected by anemia [3]. In 2021, Zambia had one of the highest burdens of anemia, ranking among the top three countries with a prevalence greater than 50% for all populations [4, 5]. Despite various interventions, the prevalence of anemia remains high, with significant disparities across different regions and demographic groups. Women of reproductive age (WRA) are at high risk due to their unique physiological processes, including menstruation, pregnancy, and childbirth. Insufficient intake of essential micronutrients like iron, vitamin B12, and folate further increases the risk.

Anemia, which significantly impacts WRA, leads to poor cognition, physical functioning, and overall quality of life, while also causing fatigue, anxiety, and depression [6]. Approximately 20% of maternal deaths are attributed to anemia (Soda et al., 2024), and during pregnancy, anemia is linked to severe outcomes such as miscarriage, early delivery, and low birth weight [7, 8]. Additionally, anemia can cause maternal mortality and other adverse neonatal outcomes (Zavala et al., 2023). In Zambia, a country with a significant proportion of women of reproductive age, anemia remains a prevalent public health problem. The Zambia Demographic and Health Survey (ZDHS) 2018 reported a national prevalence of 31% among WRA, underscoring its critical public health importance.

Despite ongoing efforts outlined in Sustainable Development Goal 3 (SDG3), anemia among WRA remains a significant global challenge. While several interventions are being put in place for the betterment of maternal and child health, high rates of anemia are ever-present, indicating the need for an understanding of its determinants and geographical distribution. [11, 12]. The prevalence of anemia can vary significantly between and within regions due to socio-economic disparities, environmental factors, healthcare access, and cultural beliefs. [10, 13].

Previous studies in Zambia, including [14], have modeled anemia as a dichotomous outcome (anemic vs non-anemic), potentially masking the differences in severity levels. Additionally, the lack of research on the geographical variations and determinants of anemia in WRA in Zambia creates a critical knowledge gap that restricts targeted intervention strategies. In contrast, the present study adopts a multilevel ordinal modeling technique for better understanding determinants of anemia severity, and spatial analysis to assess the spatial distribution, clustering of anemia and how patterns evolve over time.

This study aims to assess the spatial patterns and determine the individual, household, and community-level factors associated with anemia severity in Zambia over six years (2018 to 2024). Therefore, the contribution of this study would be the identification of the key risk factors and geographic hotspots of anemia, providing valuable insights for policymakers and public health practitioners. The findings would not only highlight the areas with the highest burden of anemia but also elucidate the underlying causes contributing to these regional disparities. Such detailed knowledge is crucial for optimizing resource allocation, designing targeted health programs, and ultimately reducing the prevalence of anemia among WRA. This study contributes significantly to the scientific literature on anemia and supports global health efforts to mitigate the impacts of anemia on vulnerable populations.

## Materials and methods

### Data

This study utilized secondary data from the Demographic and Health Surveys (DHS-https://dhsprogram.com/) Program. The DHS data are publicly available, anonymized, and contain no personally identifiable information. The permission to access the ZDHS data repository was granted on the 10^th^ of June 2024, the 2018 dataset was obtained on the 8^th^ March 2025 and the 2024 dataset was accessed on the 19^th^ November 2025. The original DHS surveys received ethical approval from the relevant national ethics committees, and informed consent was obtained from all participants before data collection. For this study, ethical clearance was obtained from the University of Zambia Biomedical Research Ethics Committee. In addition, authorization to conduct the research was granted by the National Health Research Authority in accordance with national research regulations. Further, no attempts were made to identify the subjects to protect their confidentiality.

We adopted an analytical cross-sectional study as described by Wang & Cheng (2020). The ZDHS is a nationwide representative survey conducted periodically to collect data on a wide range of demographic, health, anthropometric, and social indicators. It is part of the international Demographic and Health Surveys (DHS) program, which is coordinated by the United States Agency for International Development (USAID) and implemented by various national agencies in participating countries. The sample included all women who participated in the ZDHS (2018 and 2024) aged between 15 and 49, who provided informed consent. The study excluded everyone missing critical information required for the study.

### Sampling and data measurement

The ZDHS uses a two-stage cluster sampling design to ensure national representativeness. Clusters are selected based on probability proportional to size (PPS), and households within clusters are systematically sampled [16]. For our study, **total enumeration** was used, including all eligible women who participated in the 2018 and 2024 ZDHS surveys. Data collection instruments capture individual, household, and community-level factors. Key components include Hemoglobin levels (used to determine anemia status), demographic and socio-economic characteristics, and geographical and environmental data, including GPS coordinates. The ZDHS data for 2018 and 2024 were collected by the Zambia Statistics Agency under the standardized DHS protocol. For anemia, the blood specimens for anemia testing were collected from women aged 15-49 who consented. Blood samples were drawn from a drop of blood taken from a finger prick and collected in a microcuvette. Hemoglobin analysis was conducted using a battery-operated HemoCue® 201+ device. For our study, all datasets were accessed from the DHS program website (https://dhsprogram.com/).

### Outcome variable

The primary response variable in this study is anemia severity, which is defined based on hemoglobin levels in the blood. Which is an ordered categorical variable categorized as none, mild, moderate, and severe anemia (WHO, 2011). The WHO Hb cut-off points for anemia diagnosis are given in Table 1.

**Table 1.** Description of the anemia severity diagnosis.

| Anemia Levels | Hemoglobin level in g/L |  |
| --- | --- | --- |
|  | Pregnant women | Non-pregnant women |
| **Non-anemic** | $\geq 110$ | $\geq 120$ |
| **Mild** | 100 – 109 | 110-119 |
| **Moderate** | 70-99 | 80-109 |
| **Severe** | $\leq 70$ | $\leq 80$ |

### Data analysis

#### Descriptive Statistics

The data was summarized using descriptive statistics. Frequencies and percentages were presented. To test the association between anemia and categorical variables, the chi-square test (with design-corrected statistics accounting for clustering and stratification) was used.

#### Inferential Statistics

The multilevel (three-level) ordinal logistic regression approach was used due to the hierarchical nature of the data, that is, individuals nested in households and communities [18–20]. For all the models, we used an investigator-led backward elimination to select the best-fit model, where all candidate variables were included in the model. The AIC and BIC statistics were the primary performance metrics used in our modeling process. The adjusted odds ratios, the corresponding 95% confidence interval, and p-values were reported for the best-fit model. The proportion of variations in odds of anemia between households and clusters was expressed using the variance partition coefficient (VPC). This measures the proportion of the outcome (anemia) variation explained by the unobserved heterogeneity between subjects at each level of the model (Leckie & French, 2013). The intra-class correlation coefficients (ICCs) were calculated to measure the correlation (homogeneity) of the observed anemia cases within the households and clusters (Leckie & French, 2013).

The three-level ordinal logistic regression model [18] is used to model the ordinal anemia severity. The model is based on the cumulative probability *Y*_*ijk*_ for the *i* ― *th* individual in the *j* ― *th* household (level-2 cluster) and *k* ― *th* community (level-3 cluster) of being in category c or lower. The cumulative probability is defined as:

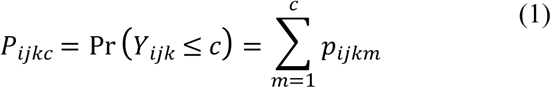

Where *p*_*ijkm*_ is the probability that an individual is in the specific anemia category *m*.

The model is specified for each cumulative logit as:

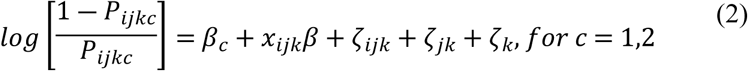

where *β*_*c*_ is the intercept for the category *c*, *c* = 3 ― 1 = 2, *β* is the fixed effect of explanatory variables, *x*_*ijk*_ is the covariate vector for the *i* ― *th* individual in the *j* ― *th* household (level-2 cluster), and *k* ― *th* community (level-3 cluster), *ζ*_*jk*_ is the level-2 random effect with variance *σ*^2^_*ζ*(2)_ and *ζ*_*k*_ is the level-3 random effect with variance *σ*^2^ _*ζ*(3)_.

Equation (2) above models the log-odds of being above category c (worse anemia) versus being in category c or below. Thus, a positive coefficient indicates that a unit increase in the explanatory variable is associated with higher log-odds of being in a more severe anemia category. Both the level-2 and level-3 random effects are assumed to be normally distributed. The individual-level variance is fixed at *π*^2^/3, just like any logistic regression model [18].

With the following key assumptions:

1. *ζ*_*jk*_ is normally distributed, independent, and has constant variance, *ζ*_*jk*_ ∽ *N*(0, *σ*^2^_*ζ*(2)_).
2. *ζ*_*k*_ is normally distributed and not correlated with covariates, *ζ*_*k*_ ∽ *N*(0, *σ*^2^_*ζ*(3)_).
3. The fixed effect represents the population-average relationship.

The model specification can be expressed mathematically as,

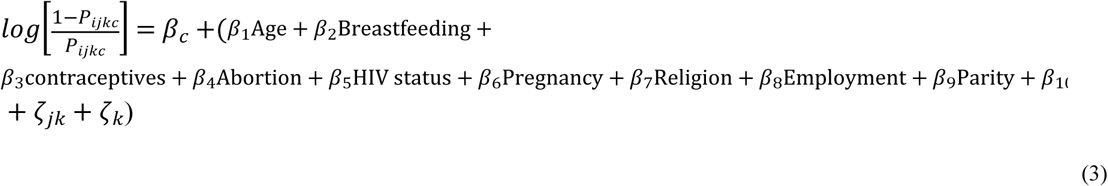

Using equation (3), we estimated a total of five (5) random intercept models. The first model (Model RI1) is the null model (the model without covariates). The second model (Model RI2) contained individual-level covariates only. The third and fourth models (Model RI3 and Model RI4) were estimated using household and community-level predictors, respectively. The last model (Model RI5) is the best fit model containing predictors at all three levels (individual, household, and community-levels).

### Variance partition coefficients (VPCs)

The variance partition coefficients (VPCs) were calculated to quantify the percentage of the variance attributable to each level of the model. The total variance in the ordinal outcome under the logistic distribution is given as the sum of the household-level variance, the community-level variance, and the level-1 (individual) variance, which is a constant *π*^2^/3. The proportion of variation in anemia severity due to the household-level random effect was calculated as the household-level variance divided by the total model variance. This represents the proportion of variance due to unobserved factors at household-level. The proportion of variation in anemia severity due to community random effects and the proportion of variation due to household and community-level random effects were obtained in the similar manner.

### Spatial analysis

Spatial data (geographic coordinates and other georeferenced data) were collected from the 2018 and 2024 Zambia demographic and health survey (ZDHS). Provincial and district administrative boundary base files were obtained from the Global Administrative Areas (GADM) database. The ZDHS data was merged with the geographic positioning system (GPS) dataset (spatial data). Anemia occurrence (proportions) was then aggregated to community (cluster), district, and provincial levels to facilitate multi-scale analysis. Spatial analyses were performed in R (R Core team, 2024).

#### Spatial autocorrelation

The Global Moran’s I, with a score between -1 and 1, was computed to assess spatial autocorrelation. Values close to 1 indicate strong positive spatial autocorrelation, meaning similar values are clustered together (high values near high values and vice versa). Values close to 0 imply the occurrence of the outcome is random, and values close to -1 indicate dispersion (dissimilar values, high values near low values, and vice versa) of the outcome occurrence. The Local Moran’s I and local Getis-Ord Gi and Gi* statistics were used to identify the hot and cold spots of anemia in women of reproductive age using GPS latitude and longitude coordinate readings taken at the nearest community center for EAs or ZDHS 2018 and 2024 clusters. The statistical significance of autocorrelation was determined by the normal distribution (z-scores) and p-values with a confidence interval of 95% (α = 0.05). Maps were used to display the distribution and variation of anemia prevalence among women of reproductive age in Zambia. The Global Moran’s I is given as [22],

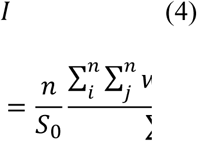

where, *n* the number of observations, *x̄* is the mean of the observations of the variable *x*, *x*_*i*_ the observations of the variable *x* at location *i*, *x*_*j*_ the observation of *x* at location *j*, *w*_*ij*_ the elements of the weight matrix, *S*_0_ The sum of the elements of the weight matrix.

#### Spatial scan statistics

Spatial scan statistics, for the two waves (2018 and 2024), were employed to identify Primary and secondary clusters of anemia using R v 5.2.1 (using Kuldorff’s SatScan package). To fit the Bernoulli model, the response variable (anemia severity) was dichotomized into anemic (mild, moderate and severe) and non-anemic. Women categorized as anemic were further classified as cases and the non-anemic as controls. In 2018 a total of 535 (10 clusters excluded due to implausible coordinates) enumeration areas were used and 545 for the 2024 cycle. The upper limit for the scanning window was the default maximum spatial cluster size of less than 50% of the total population at risk. This allows the detection of both small and large clusters while excluding clusters with the majority of the study population. Areas with significant p-values (estimated through 999 Monte Carlo replications) were considered as high anemic areas compared to areas outside the window.Despite the cross-sectional nature of the ZDHS, we compared spatial patterns across waves allowing us to assess the temporal changes in the spatial distribution, providing a pseudo-spatial temporal perspective.

### Ethical Consideration

The study utilizes secondary data from the Zambia Demographic and Health Survey (ZDHS), and permission to access the data was granted by https://dhsprogram.com/data. Since ZDHS data is georeferenced, it enables spatial analysis to be conducted as outlined in our methodology. To maintain the confidentiality of respondents, ZDHS has a georeferenced data release policy. The policy provides two levels of protection. First, data from the same enumeration area (EA) are aggregated to a single-point coordinate. Then, ZDHS applies a GPS coordinate displacement process, which masks the true locations: urban clusters are displaced a distance of up to two kilometers (0-2 km), rural clusters up to five kilometers (0-5 km), and an additional randomly selected 1% of rural clusters are displaced up to ten kilometers (0-10 km) [23]. This study upholds all ZDHS confidentiality guidelines and data-sharing policies to ensure ethical compliance.

## Results

### Descriptive statistics

The study included a total of 19,362 women of reproductive age, of which 12,735 (65.77%) were from the 2018 survey and 6,627 (34.23%) from 2024 survey. The overall prevalence of mild, moderate and severe anemia was 16.3% (3,148), 13.2% (2,545) and 1.4% (268), respectively.

The prevalence of anemia in 2018 was 31.0% (n=3,946), with a 95% confidence interval (95% CI) of 29.5% to 32.4%. In 2024 the estimated prevalence was 30.4% (n=2,015), with a 95% confidence interval of 29.1% to 31.7%. The difference in the prevalence of anemia between the year 2018 and 2024 was not statistically significant (p=0.559). Similarly, there was no evidence of an association between anemia severity and survey year (2018 and 2024) (see Table 2).

**Table 2.** Sociodemographic and health-related characteristics of women of reproductive age by anemia severity.

|  | **Anemia Severity** |  |  |  |  |  |
| --- | --- | --- | --- | --- | --- | --- |
| **Variable** | **Non-Anemic** | **Mild** | **Moderate** | **Severe** | **Total (Weighted %)** | **P-value** |
| **Survey Year** |  |  |  |  |  |  |
| 2018 | 8788 | 2065 | 1710 | 172 | 12,735 (65.77%) | 0.647 |
| 2024 | 4612 | 1083 | 836 | 96 | 6,627 (34.23%) |  |
| **Age group** |  |  |  |  |  |  |
| 15-19 | 2884 | 833 | 615 | 63 | 4,394 (22.70%) | <0.001 |
| 20-24 | 2674 | 561 | 451 | 38 | 3,725 (19.24%) |  |
| 25-29 | 2288 | 511 | 336 | 32 | 3,168 (16.36%) |  |
| 30-34 | 1905 | 391 | 322 | 36 | 2,654 (13.71%) |  |
| 35-39 | 1580 | 367 | 330 | 32 | 2,310 (11.93%) |  |
| 40-44 | 1205 | 281 | 293 | 44 | 1,823 (9.41%) |  |
| 45-49 | 864 | 204 | 198 | 22 | 1,288 (6.65%) |  |
| **HIV status** |  |  |  |  |  |  |
| Negative | 11965 | 2688 | 1982 | 182 | 16,818 (86.86%) | <0.001 |
| Positive | 1436 | 460 | 563 | 86 | 2,544 (13.14%) |  |
| **Marital status** |  |  |  |  |  |  |
| Never in union | 4086 | 1117 | 896 | 114 | 6,213 (32.09%) | <0.001 |
| Currently in union | 7679 | 1596 | 1254 | 120 | 10,649 (55.00%) |  |
| Formerly in union | 1635 | 435 | 395 | 34 | 2,499 (12.91%) |  |
| **Employment** |  |  |  |  |  |  |
| No | 7226 | 1749 | 1403 | 156 | 10,534 (54.40%) | 0.400 |
| Yes | 6174 | 1399 | 1143 | 112 | 8,828 (45.60%) |  |
| **Pregnancy** |  |  |  |  |  |  |
| No | 12512 | 2838 | 2280 | 254 | 17,884 (92.37%) | <0.001 |
| Yes | 888 | 310 | 266 | 14 | 1,478 (7.63%) |  |
| **Contraceptive use** |  |  |  |  |  |  |
| No contraceptive use | 7926 | 2158 | 1827 | 205 | 12,116 (62.58%) | <0.001 |
| Using any method | 5475 | 990 | 718 | 63 | 7,245 (37.42%) |  |
| **Breastfeeding** |  |  |  |  |  |  |
| No | 10380 | 2456 | 2103 | 240 | 15,180 (78.40%) | <0.001 |
| Yes | 3020 | 691 | 442 | 28 | 4,182 (21.60%) |  |
| **Parity** |  |  |  |  |  |  |
| Nulliparous | 3201 | 913 | 723 | 104 | 4,941 (25.52%) | <0.001 |
| Primiparous | 2205 | 519 | 460 | 46 | 3,230 (16.68%) |  |
| Multiparous | 4815 | 1022 | 820 | 61 | 6,718 (34.70%) |  |
| **Gravidity** |  |  |  |  |  |  |
| 0 | 3201 | 913 | 723 | 104 | 4,941 (25.52%) | <0.001 |
| 1--3 | 5708 | 1240 | 1046 | 84 | 8,078 (41.72%) |  |
| >=4 | 4491 | 995 | 776 | 80 | 6,343 (32.76%) |  |
| **Children in past 5yrs** |  |  |  |  |  |  |
| None | 6165 | 1548 | 1403 | 193 | 9,309 (48.08%) | <0.001 |
| One | 5134 | 1099 | 831 | 64 | 7,128 (36.82%) |  |
| Two and More | 2101 | 500 | 311 | 11 | 2,924 (15.10%) |  |
| **Abortion** |  |  |  |  |  |  |
| No | 12058 | 2858 | 2278 | 225 | 17,419 (89.97%) | 0.001 |
| Yes | 1343 | 290 | 267 | 43 | 1,942 (10.03%) |  |
| **Religion** |  |  |  |  |  |  |
| Catholic | 2220 | 498 | 426 | 35 | 3,179 (16.42%) | 0.095 |
| Protestant | 11023 | 2597 | 2072 | 230 | 15,922 (82.24%) |  |
| Muslim | 65 | 14 | 9 | 0 | 88 (0.45%) |  |
| Other | 93 | 38 | 38 | 3 | 172 (0.89%) |  |
| **Smoking status** |  |  |  |  | 0.00% |  |
| No | 13031 | 3065 | 2484 | 268 | 18848 (97.35%) | 0.038 |
| Yes | 369 | 83 | 61 | 0 | 513 (2.65%) |  |
| **Owns net** |  |  |  |  |  |  |
| No | 2422 | 598 | 505 | 58 | 3582 (18.50%) | 0.29 |
| Yes | 10979 | 2550 | 2041 | 210 | 15779 (81.50%) |  |
| Others | 78 | 10 | 19 | 3 | 110 (0.57%) |  |
| **Wealth index** |  |  |  |  |  |  |
| Poorest | 2386 | 589 | 444 | 25 | 3445 (17.79%) | <0.001 |
| Poorer | 2349 | 625 | 403 | 37 | 3415 (17.64%) |  |
| Middle | 2511 | 597 | 440 | 49 | 3597 (18.58%) |  |
| Richer | 3029 | 622 | 588 | 78 | 4317 (22.30%) |  |
| Richest | 3125 | 715 | 669 | 79 | 4587 (23.69%) |  |
| **Money for medical help** |  |  |  |  |  |  |
| Big problem | 2785 | 677 | 613 | 58 | 4133 (21.35%) | 0.032 |
| Not a big problem | 10615 | 2471 | 1932 | 211 | 15229 (78.65%) |  |
| **Family size** |  |  |  |  |  |  |
| Less than 3 | 691 | 172 | 188 | 18 | 1069 (5.52%) | 0.006 |
| 3 and 4 | 3288 | 715 | 591 | 57 | 4651 (24.02%) |  |
| More 4 | 9421 | 2261 | 1766 | 193 | 13642 (70.46%) |  |
| **Sex HH** |  |  |  |  | 0.00% |  |
| Male | 9619 | 2223 | 1706 | 186 | 13733 (70.93%) | 0.001 |
| Female | 3782 | 925 | 840 | 82 | 5628 (29.07%) |  |
| **Residency** |  |  |  |  |  |  |
| Urban | 6358 | 1392 | 1301 | 153 | 9204 (47.54%) | <0.001 |
| Rural | 7043 | 1756 | 1245 | 115 | 10158 (52.46%) |  |
| **Province** |  |  |  |  |  |  |
| Central | 1403 | 265 | 185 | 19 | 1872 (9.67%) | <0.001 |
| Copperbelt | 2096 | 451 | 356 | 42 | 2944 (15.21%) |  |
| Eastern | 1645 | 367 | 239 | 20 | 2271 (11.73%) |  |
| Luapula | 970 | 265 | 186 | 15 | 1435 (7.41%) |  |
| Lusaka | 2536 | 598 | 621 | 79 | 3834 (19.80%) |  |
| Muchinga | 720 | 154 | 110 | 7 | 992 (5.12%) |  |
| Northern | 1111 | 252 | 156 | 8 | 1527 (7.89%) |  |
| North-Western | 712 | 214 | 163 | 18 | 1108 (5.72%) |  |
| Southern | 1499 | 391 | 317 | 40 | 2247 (11.61%) |  |
| Western | 709 | 190 | 213 | 20 | 1131 (5.84%) |  |
| **Total** | **13,400**<br>**(69.21%)** | **3,148**<br>**(16.26%)** | **2,545**<br>**(13.15%)** | **268**<br>**(1.38%)** | **19,362 (100.00%)** |  |

All p-values obtained using Chi-square test for independence adjusting the for weights and sampling design.

Over 40% (8,119) of the women were adolescents and young adults aged 15 to 24 years, and age was significantly associated with anemia severity. Of all the participants, 2,544 (13.14%) were HIV positive, and HIV status showed a significant association with anemia severity. Reproductive and obstetric characteristics including pregnancy status, breastfeeding (yes/no), contraceptive, parity, gravidity, children born in the past five years and history of abortion were all strongly associated with anemia severity (see Table 2).

Household and community-level determinants equally showed significant associations with anemia severity. Wealth index was associated with anemia severity (p<0.001), with over 30% (6,860) of the women belonging to either the poorer or poorest. Over 50% (10,158) of the women of reproductive age in our study had rural residency, and residency (rural/urban) was strongly associated with anemia severity. Finally, there was clear of sub-national variation in anemia severity (p<0.001), with provinces showing strong associated with anemia severity. Which amplifies the existing geographical disparities.

### Multivariable multilevel ordinal logistic regression

#### Random Effects

The intercept-only model (null model) is the model without independent variables. It provided the variance components of random effects between household and community clusters to justify multilevel analysis over standard models. The total percentage of variance in anemia severity due to household and community heterogeneity combined was 37.14% (*VPC*_2+3_). Of this total variation, 1.39% (*VPC*_3_) was attributed to unobserved community-level factors, while 35.75% was due to unobserved household-level factors. These variance proportion coefficients indicate dependency at the household and community level, which justifies multilevel modelling (Table 3).

**Table 3.** Random intercept variances and model fit statistics of three-level mixed effect models.

| Random effects | Null Model | Model RI5 |
| --- | --- | --- |
| $\sigma^2_{\zeta(3)}$ | 0.073 | 0.052 |
| $\sigma^2_{\zeta(2)}$ | 1.871 | 1.862 |
| $VPC_2\%$ | 35.748 | 35.767 |
| $VPC_3\%$ | 1.395 | 0.999 |
| $VPC_{2+3}\%$ | 37.142 | 36.780 |
| $PCV_3$ | | 28.767 |
| **Model fit stats** |  |  |
| **AIC** | 31,130.94 | 30572.18 |
| **BIC** | 31,162.42 | 30,800.39 |

From the best fit Model (Model RI5),it was observed that 36.78% (*VPC*_2+3_), 1.00% (*VPC*_3_), and 35.78% (*VPC*_2_) of the unexplained variation in anemia could be attributed to unobserved community and household level factors combined, community level factors and household level factors, respectively. The *PCV*_3_showed that 28.77% of the community-level variation was explained by the variables in the model. This suggests that, after controlling for variables in the model, there are still significant factors of anemia at household and community levels which are not accounted for in the model. The best fit model, Model RI5 was selected based on the BIC and AIC statistics (see Table 3).

#### Factors Associated with Anemia Severity

Table 4 below (Model RI5) presents factors associated with anemia among WRA. The same odds ratios were interpreted comparing higher versus lower levels of anemia (moderate/severe versus mild and non-anemic, mild versus non-anemic). Three variables, age, births in the past five years and residency, violated the proportional odds assumption based on the Brant test. Nonetheless, the proportional odds model was retained due to the principle of parsimony, superior overall fit, and ease of interpretation. The best-fit model summarized the associations between anemia severity and the predictors, estimating the overall direction and magnitude of effects, contrary to category-specific contrasts.

**Table 4.** Factors associated with anemia severity among women of reproductive age in Zambia, Data from ZDHS (2018 and 2024, n=19,362)

|  | **Unadjusted Estimates** |  | **Adjusted Estimates** |  |
| --- | --- | --- | --- | --- |
| **Variable** | **COR [95% CI]** | **P-value** | **AOR [95% CI]** | **P-value** |
| **Survey Year** |  |  |  |  |
| 2018 | Reference | n/a | Reference | n/a |
| 2024 | 0.997 [0.900, 1.105] | 0.954 | 1.038 [0.939, 1.148] | 0.467 |
| **Age group** |  |  |  |  |
| 15-19 | Reference | n/a | Reference | n/a |
| 20-24 | 0.716 [0.624, 0.823] | <0.001 | 0.830 [0.711, 0.968] | 0.018 |
| 25-29 | 0.645 [0.557, 0.747] | <0.001 | 0.786 [0.657, 0.941] | 0.009 |
| 30-34 | 0.731 [0.621, 0.860] | <0.001 | 0.866 [0.710, 1.057] | 0.157 |
| 35-39 | 0.853 [0.726, 1.001] | 0.052 | 1.025 [0.836, 1.257] | 0.813 |
| 40-44 | 1.042 [0.875, 1.241] | 0.646 | 1.192 [0.947, 1.499] | 0.135 |
| 45-49 | 0.977 [0.820, 1.164] | 0.795 | 1.075 [0.863, 1.338] | 0.521 |
| **HIV status** |  |  |  |  |
| Negative | Reference | n/a | Reference | n/a |
| Positive | 2.519 [2.156, 2.943] | <0.001 | 2.632 [2.246, 3.085] | <0.001 |
| **Marital status** |  |  |  |  |
| Never in union | Reference | n/a | Reference | n/a |
| Currently in union | 0.700 [0.635, 0.772] | <0.001 | 0.663 [0.572, 0.769] | <0.001 |
| Formerly in union | 1.109 [0.961, 1.280] | 0.158 | 0.883 [0.730, 1.067] | 0.198 |
| **Employment** |  |  |  |  |
| No |  |  | Reference | n/a |
| Yes | 0.891 [0.811, 0.979] | 0.016 | 0.912 [0.826, 1.006] | 0.067 |
| **Pregnancy** |  |  |  |  |
| No | Reference | n/a | Reference | n/a |
| Yes | 1.612 [1.382, 1.880] | <0.001 | 1.961 [1.665, 2.310] | <0.001 |
| **Births in past 5yrs** |  |  |  |  |
| None | Reference | n/a | Reference | n/a |
| One | 0.675 [0.606, 0.752] | <0.001 | 0.842 [0.741, 0.956] | 0.008 |
| Two and More | 0.678 [0.590, 0.779] | <0.001 | 0.960 [0.821, 1.123] | 0.609 |
| **Money for medical help** |  |  |  |  |
| Big problem | Reference | n/a | Reference | n/a |
| Not a big problem | 0.816 [0.731, 0.911] | <0.001 | 0.847 [0.757, 0.948] | 0.004 |
| **Residency** |  |  |  |  |
| Urban | Reference | n/a | Reference | n/a |
| Rural | 1.025 [0.920, 1.142] | 0.653 | 1.210 [1.080, 1.354] | 0.001 |
| **Province** |  |  |  |  |
| Central | Reference | n/a | Reference | n/a |
| Copperbelt | 1.201 [0.980, 1.471] | 0.077 | 1.260 [1.021, 1.555] | 0.031 |
| Eastern | 1.116 [0.907, 1.374] | 0.300 | 1.171 [0.952, 1.440] | 0.134 |
| Luapula | 1.547 [1.272, 1.881] | <0.001 | 1.650 [1.355, 2.010] | <0.001 |
| Lusaka | 1.677 [1.358, 2.072] | <0.001 | 1.804 [1.457, 2.233] | <0.001 |
| Muchinga | 1.151 [0.857, 1.544] | 0.350 | 1.236 [0.916, 1.669] | 0.166 |
| Northern | 1.091 [0.854, 1.393] | 0.485 | 1.194 [0.933, 1.528] | 0.158 |
| North_Western | 1.875 [1.483, 2.370] | <0.001 | 2.003 [1.570, 2.555] | <0.001 |
| Southern | 1.517 [1.240, 1.856] | <0.001 | 1.500 [1.224, 1.838] | <0.001 |
| Western | 2.239 [1.790, 2.800] | <0.001 | 2.097 [1.676, 2.624] | <0.001 |
| **Cut1** |  |  | 1.254 [1.040, 1.468] |  |
| **Cut2** |  |  | 2.550 [2.296, 2.803] |  |
| **AIC** |  |  | 30, 572.18 |  |
| **BIC** |  |  | 30, 800.39 |  |

Models adjusted for all variables listed in the table and survey design using a three-level mixed-effects ordinal logistics regression model. Reference categories are indicated as reference in the table. Crude odds ratios (COR), adjusted odds ratio (AOR) and Confidence interval (CI).

The odds of being in a higher category of anemia for women in the 2024 survey were 1.04 times greater than the odds of women from the 2018 survey. However, we could not rule out chance finding as there was no evidence of a difference in anemia severity between the years. The odds of being above a particular category of anemia for HIV positive women were 2.63 times the odds for HIV negative women, keeping the other predictors constant (AOR=2.63, 95% CI: 2.25, 3.09). Pregnant women had 1.96 times greater odds of being in a higher category of anemia compared to non-pregnant women, adjusting for the other covariates. On the other hand, having one birth in the past five years reduced the odds of worse anemia by 16% relative to having no birth (AOR=0.84, 95% CI: 0.74, 0.96).

Women who were currently in union had reduced the odds of anemia severity by 33.7% compared to women who had never been in a union. Similarly, women who were formerly in a union had 11.7% less odds of having higher levels of anemia compared to women who had never been in a union. However, we could not rule out chance finding as there was no evidence of an association.

Women in employment had 0.91 times lesser odds of being in a higher level of anemia compared to their unemployed counterparts. However, this effect was not statistically significant.

For age, a non-linear relationship with anemia severity was observed, women aged 35 to 39 years, 40 to 44 years, and 45 to 49 years had 1.03, 1.19- and 1.08-times higher odds of a higher level of anemia compared to women aged 15 to 19 years, respectively. While women aged 20-24 had 0.83 times lower odds of being in a higher category of anemia compared to women aged 15-19. Similarly, women aged 25-29 years had 0.79 times lower odds of anemia severity.

At the household level, women who responded that money for medical help was not a big problem had 15.3% lower odds of worse anemia compared to women who admitted it was a big problem, at 95% confidence interval the odds could be 5.2% to 24.3% lower.

At the community level, subnational variations were observed in the data. Rural residence was associated with 21% higher odds of anemia severity compared to urban areas (AOR = 1.21, 95% CI: 1.08-1.35). The odds of being in a higher category of anemia were 1.26, 1.65, 1.80, 2.00, 1.50, and 2.10 times greater for women living in the Copperbelt, Luapula, Lusaka, North-Western, Southern, and Western provinces compared to women from Central province, respectively.

To further explore the observed geographic heterogeneity in anemia severity at provincial level among women of reproductive age, we conducted spatial analyses.

### Spatial analysis

The maps in **Fig 1** show that regional variations in anemia prevalence among women of reproductive age are present both at provincial and district levels for the years 2018 and 2024. The prevalence of anemia across the provinces was significantly different (p-value = < 0.001) for both years. The spatial distribution in 2024 showed elevated prevalence in North-Western, Luapula and Western provinces. Suggesting a geographical shift of anemia towards the north from 2018 to 2024 at both district and provincial level.

**Fig 1:**
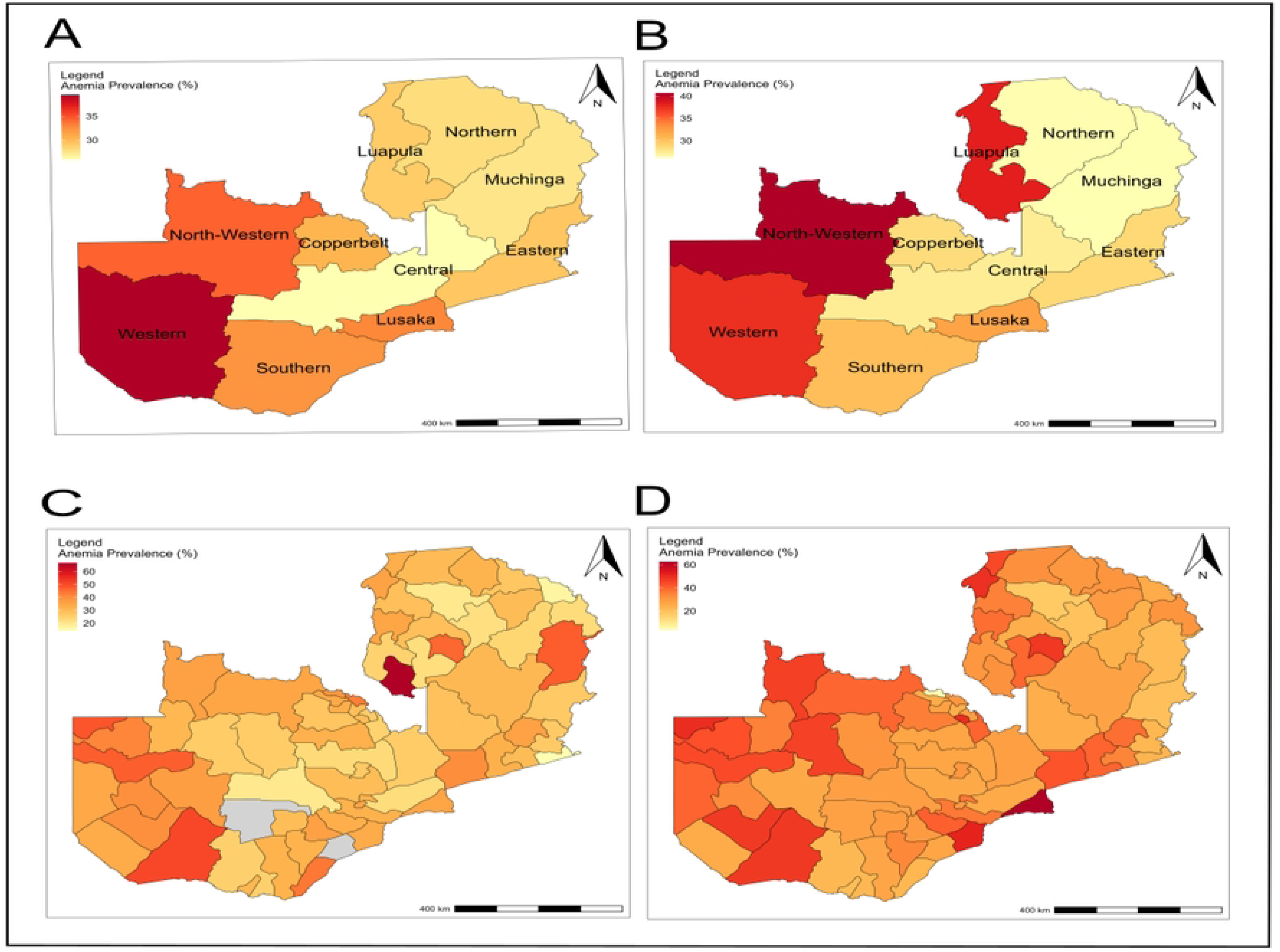
Spatial distribution of anemia among women of reproductive age in Zambia at provincial and district level in 2018 and 2024. Boundaries obtained from the GADM database of Global Administrative Areas (GADM, 2025). The color gradient (yellow to red) the prevalence distribution with dark red depicting high prevalence of anemia. (A) Anemia prevalence at provincial level 2018, (B) Anemia prevalence at provincial level 2024, (C) Anemia prevalence at district level 2018 and (D) Anemia prevalence at district level 2024

We identified the hotspot and cold-spot areas of anemia among women of reproductive age. The red color represents hotspots of anemia prevalence (high clusters), which were observed in the Western, North-Western, Southern, Copperbelt, Lusaka province, and a few other parts of the country. In contrast, blue color indicates cold spot areas (low clusters), which were predominantly observed in Central, Northern, Muchinga, and Eastern provinces (**Fig 2**) (2018). In 2024, a northward expansion of the hotspots (high risk areas) was observed, with statistically significant hotspots emerging prominently in North-Western and Luapula, which were not present in 2018.

**Fig 2:**
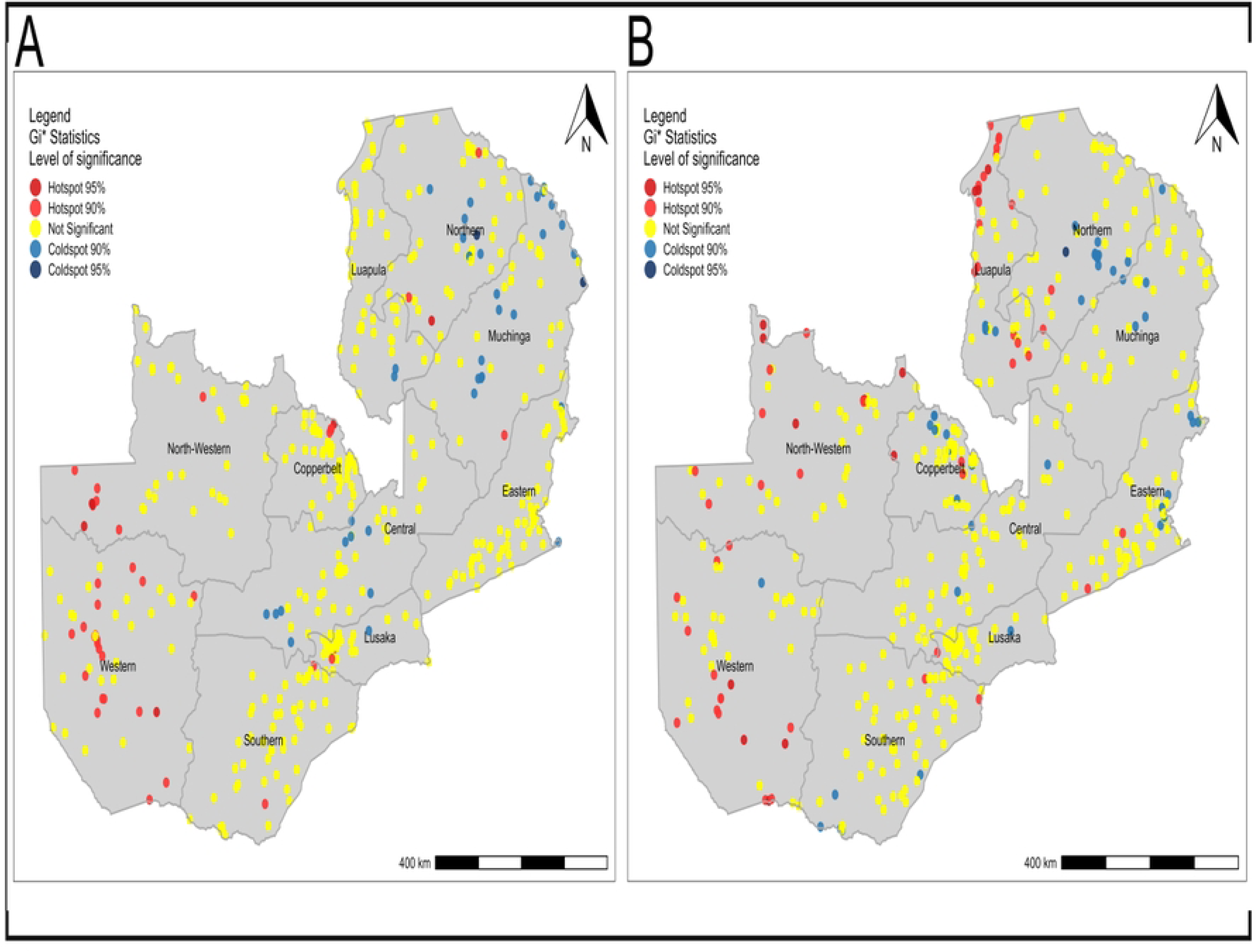
Hotspots and coldspots of anemia in WRA 2018 and 2024. Boundaries obtained from the GADM database of Global Administrative Areas (GADM, 2025). (A) Hot spot distribution of anemia 2018 and (B) Hot spot distribution of anemia 2024.

Despite the prevalence of anemia between the waves being constant (insignificant difference), the prevalence in anemia at district level was on the rise as exhibited in **Fig 3**. Very few districts (less 20) showed no or small reduction in anemia prevalence **(Fig 3)**.

**Fig 3:**
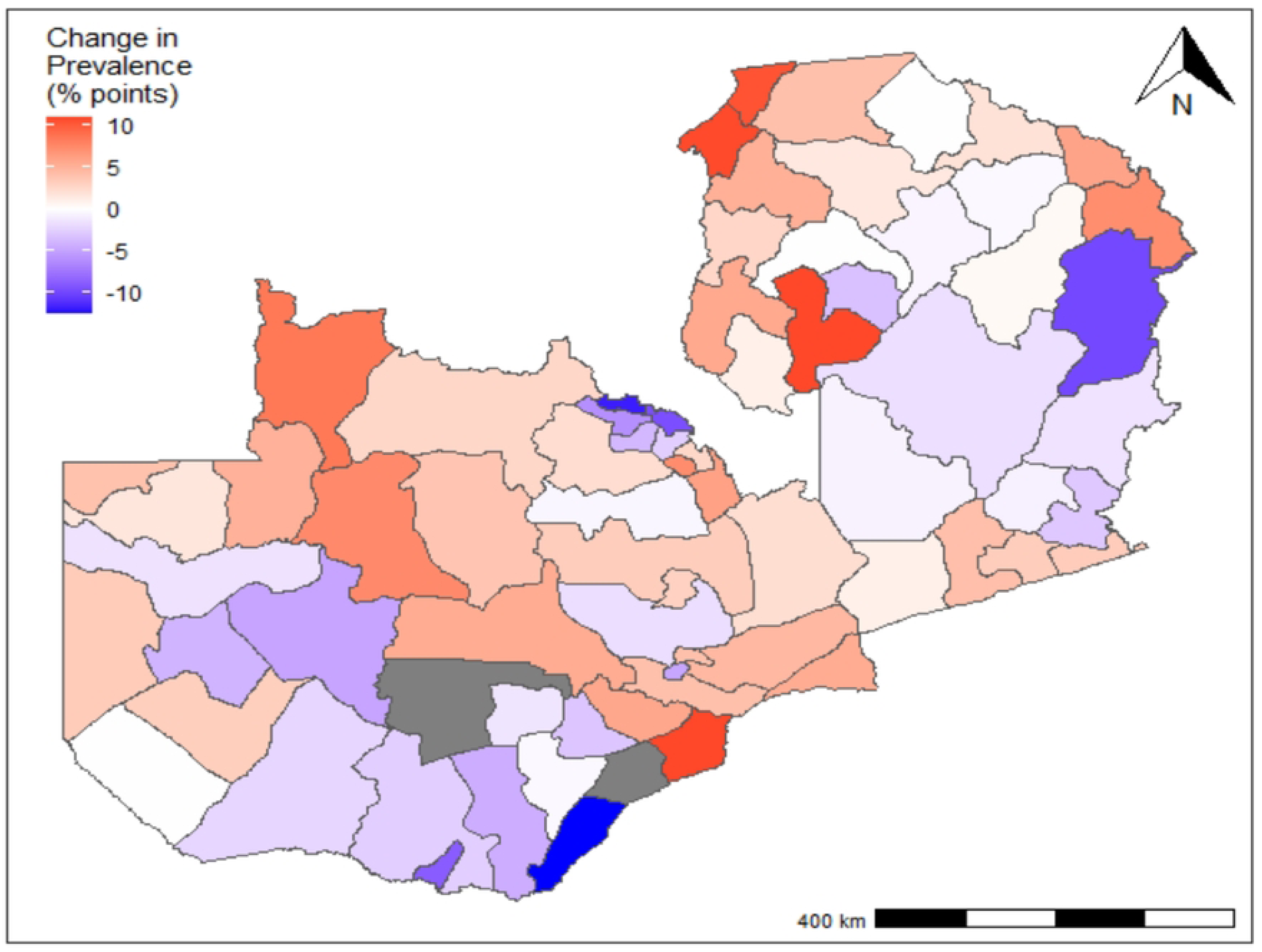
Percentage change in anemia prevalence from 2018 to 2024. Boundaries obtained from the GADM database of Global Administrative Areas (GADM, 2025). The color gradient depicts the change in anemia prevalence in the two waves, with red indicating an increase and blue a reduction.

In 2018, at the district level, the global Moran’s I statistic was 0.064, indicating that anemia prevalence was randomly distributed (not clustered; p-value 0.166). However, at the cluster level, there was some level of clustering observed with a global Moran’s I score of 0.147. Given the Z-score of 5.879, there is less than 1% probability that this pattern is due to chance (Table 5). In 2024, spatial clustering was observed at both district and cluster levels. District-level clustering was significant (Moran’s I=0.168, p=0.011), implying non-random geographic patterns. At the cluster level, spatial clustering was high (Moran’s I=0.130, z=5.202, p<0.001). The results show that anemia is spatially distributed in Zambia, with clustering more evident at finer geographic scales.

**Table 5.** Global Moran’s statistics for spatial autocorrelation of anemia prevalence.

| Level | Moran's I | E(I) | var(I) | Z(I) | p-value |
| --- | --- | --- | --- | --- | --- |
| District 2018 | 0.064 | -0.014 | 0.007 | 0.97 | 0.166 |
| Cluster 2018 | 0.147 | -0.002 | 0.001 | 5.879 | < 0.001 |
| District 2024 | 0.168 | -0.014 | 0.006 | 2.301 | 0.011 |
| Cluster 2024 | 0.130 | -0.002 | 0.001 | 5.202 | < 0.001 |

A total of 216 and 145 enumeration areas fell in the primary and secondary cluster spatial windows of anemia risk 2018 and 2024 respectively. In 2018, a total of 206 EAs were shown to lie in the primary clusters in the south-west region of the country, with the following coordinates longitude 22.453, and latitude -16.546 and a radius of 664.99 km. The relative risk corresponding to the primary cluster was 1.23 with p<0.001. This implies that women in the spatial window had 1.23 higher risk of being anemic compared to women outside the spatial window. Similarly, in 2024 the primary cluster was identified in the south-west region of Zambia, with a total of 90 EAs (Table 6 and Fig. 4). Women in the primary spatial window had an increased risk of experiencing anemia of 1.29 (RR = 1.29, p < 0.001) (Table 6). The primary cluster of anemia among women of reproductive in 2018 and 2024 was in the same region covering about four provinces (Western, North-Western, Southern and Central) with a slight reduction in the spatial window, evidence by the reduction in the radius (Table 6 and Fig. 4).

**Fig 4:**
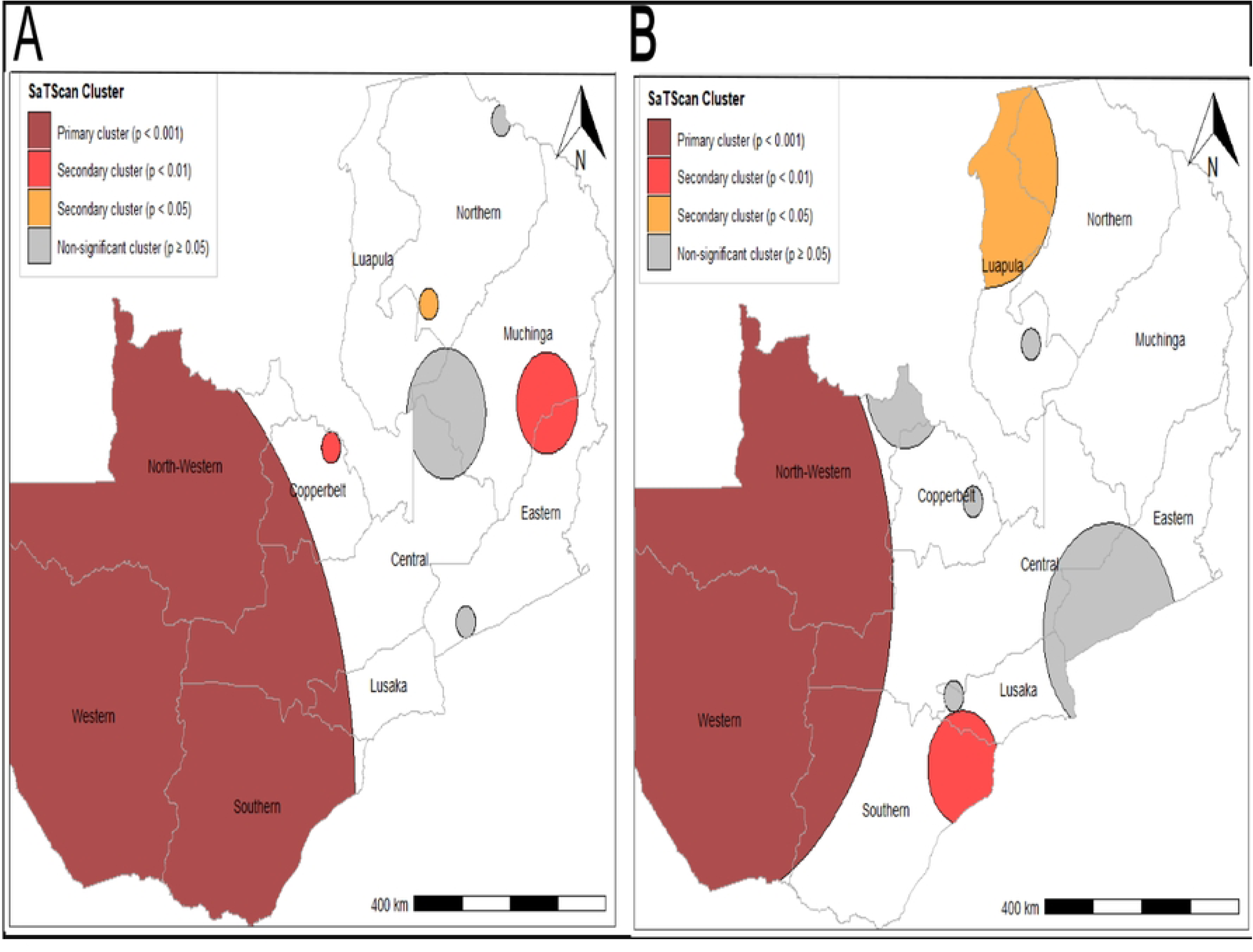
Primary and secondary clusters of WRA 2018 and 2024. Boundaries obtained from the GADM database of Global Administrative Areas (GADM, 2025). Dark red color represents the primary clusters (most likely clusters), that is the cluster with the highest likelihood ratio. While, light red and dark orange are secondary clusters from the highest to lowest the likelihood ratio. Grey represents clusters that are not statistically significant. (A) Satscan analysis for 2018 and (B) Satscan analysis for 2024.

**Table 6.**
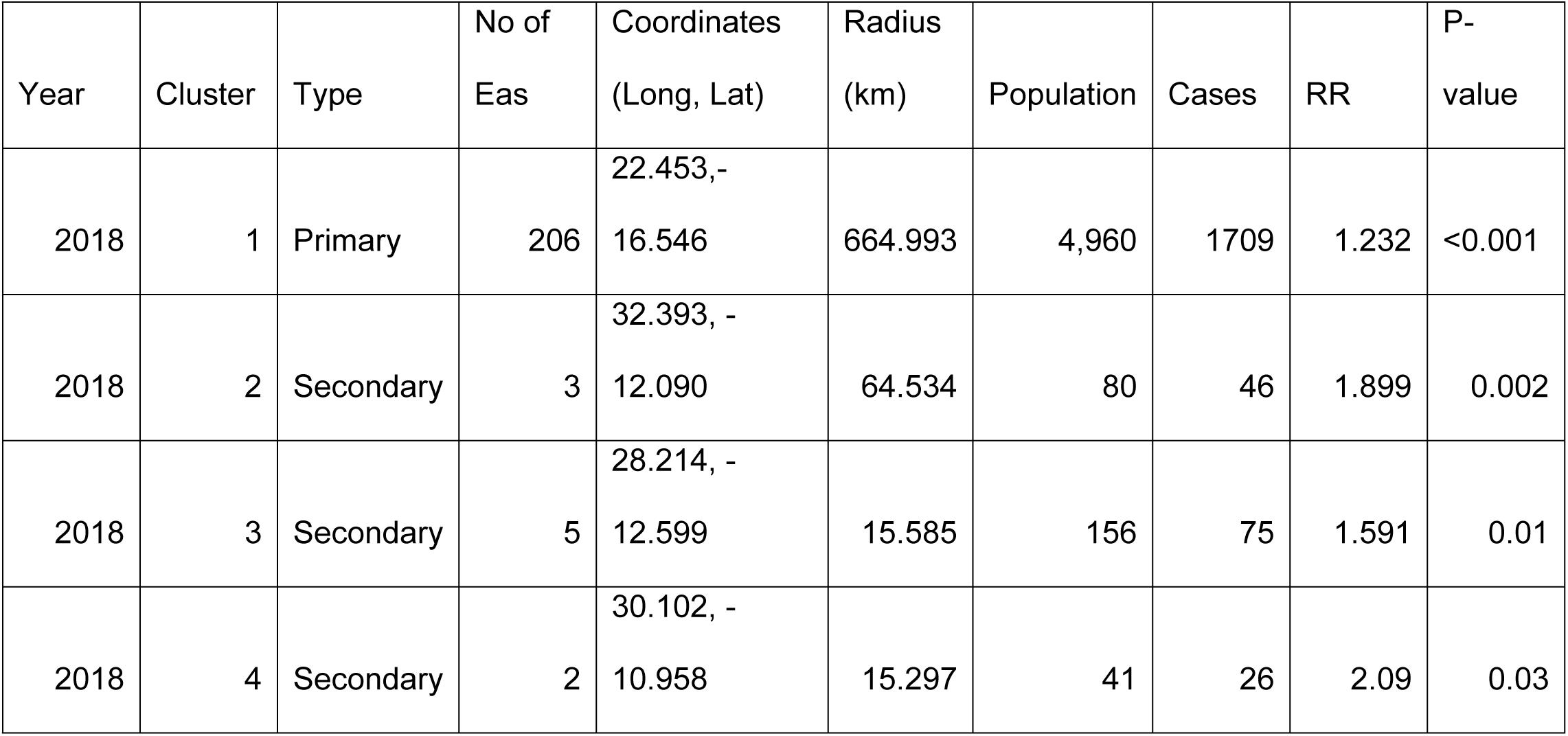
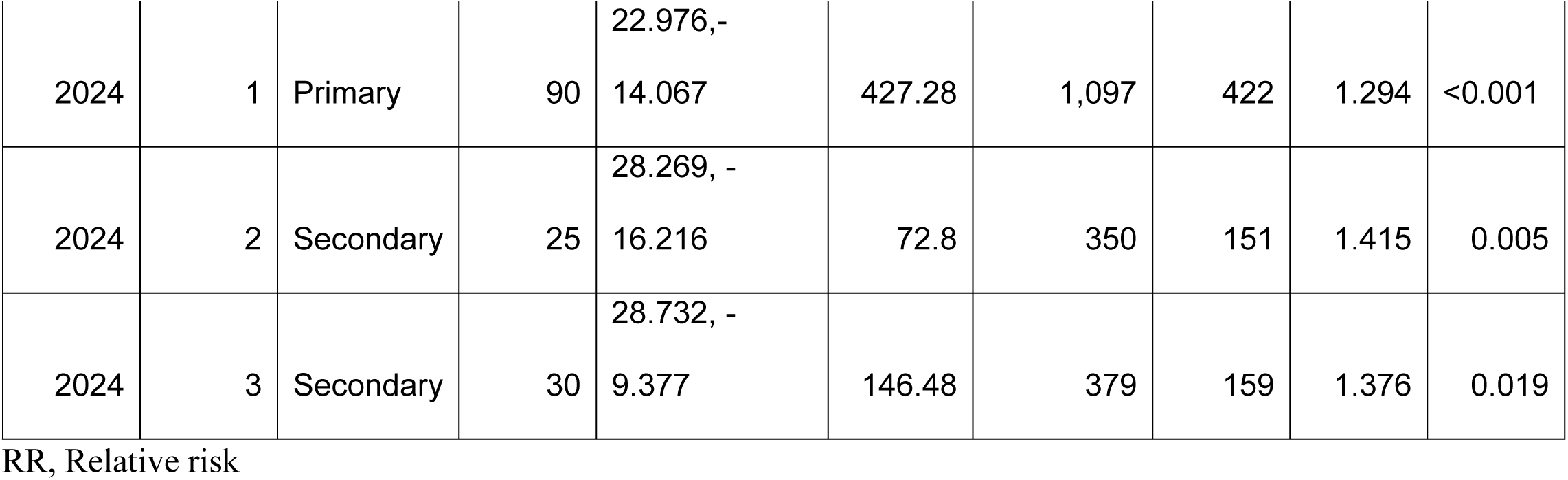
Spatial clusters of Anemia among women of reproductive age (15-49) (2018 and 2024 ZDHS)

| Year | Cluster | Type | No of<br>Eas | Coordinates<br>(Long, Lat) | Radius<br>(km) | Population | Cases | RR | P-value |
| --- | --- | --- | --- | --- | --- | --- | --- | --- | --- |
| 2018 | 1 | Primary | 206 | 22.453,-<br>16.546 | 664.993 | 4,960 | 1709 | 1.232 | <0.001 |
| 2018 | 2 | Secondary | 3 | 32.393, -<br>12.090 | 64.534 | 80 | 46 | 1.899 | 0.002 |
| 2018 | 3 | Secondary | 5 | 28.214, -<br>12.599 | 15.585 | 156 | 75 | 1.591 | 0.01 |
| 2018 | 4 | Secondary | 2 | 30.102, -<br>10.958 | 15.297 | 41 | 26 | 2.09 | 0.03 |
| 2024 | 1 | Primary | 90 | 22.976,-<br>14.067 | 427.28 | 1,097 | 422 | 1.294 | <0.001 |
| 2024 | 2 | Secondary | 25 | 28.269, -<br>16.216 | 72.8 | 350 | 151 | 1.415 | 0.005 |
| 2024 | 3 | Secondary | 30 | 28.732, -<br>9.377 | 146.48 | 379 | 159 | 1.376 | 0.019 |

RR, Relative risk

## Discussion

Our study shows a stable prevalence of anemia, with no significant difference in prevalence over the six years, 31.0% (3,946) in 2018 and 30.4% (2,015) in 2024). Therefore, anemia remains a moderate public health concern in Zambia, according to the World Health Organization (WHO) criteria. Despite stable overall anemia rates, the spatial risk distribution was dynamic, as shown by shifts in hotspot locations over time. This indicates that the underlying causes of anemia may be changing geographically. Overall, the study found that anemia is influenced by multiple factors at the individual, household, and community levels. There was significant unobserved heterogeneity at the household-level and smaller but not negligible variation at the community-level. Additionally, the lack of reduction in anemia over the six years suggests that current interventions have limited success in addressing the root causes of anemia.

### Multilevel Determinants of Anemia

Age, HIV status, pregnancy, Marital status, employment status, births in the past five years, money for medical assistance, residency and province, were significantly associated with anemia severity after adjusting for individual household-level and community-level factors.

At the individual level, HIV infection was strongly associated with anemia severity, this is consistent with previous findings [14]. The high susceptibility of HIV patients is due to several factors which include, antiretroviral-related hematologic effects, decreased red blood cells production, increased red blood cell destruction and ineffective red blood cell production, and opportunistic infections [24]. Similarly, pregnancy emerged as a strong predictor of anemia severity. This highlights the established biological mechanisms such as increased iron requirements, nutritional depletion, vaginal and parasitic infections during pregnancy [25]. Women with one birth had decreased odds of anemia severity compared to women with no birth in the past five years. This may reflect consistent antenatal care, including iron supplementation rather than a direct effect of childbirth.

Women aged 15 to 19 years had higher odds of worse anemia compared to women aged 20 to 29 years. This corresponds with findings in Ethiopia and Mali, where younger women had a higher risk [26–29]. The increased risk in adolescents could be attributed to menstrual blood loss, and early childbearing. Women categorized as currently married had decreased odds of a higher level of anemia compared to women who had never been married [25, 30, 31].

The estimated variance partition coefficient indicated that over one-third of the variance in anemia severity was due to unobserved heterogeneity at household-level. This implies that women in the same household share similar unmeasured anemia risks. Women with fewer financial constraints towards medical assistance had lower odds of anemia severity. The link between financial access and health suggests that anemia in Zambia is not solely a nutritional issue but is tied to the ability to afford high-quality care and supplements.

Despite community variance being lower than household variance, geographical variation remained after adjustments. The rural residency was associated with higher odds of anemia severity compared to urban residency, this was consistent with studies by Ara et al. (2019), Gona et al. (2021), and Mankelkl & Kinfe (2023b), highlights the existing inequality in the health care system. While some literature [35] argues that urban areas can sometimes present a higher risk due to poor diet and overpopulation, the Zambian context confirms that rural women are more vulnerable; this could be due to limited access to diversified diets and healthcare.

### Spatial distribution and regional disparities

One of the most important findings of our study is the clustered nature of anemia in Zambia. A geographical expansion of hotspots was observed, from Western to North-Western and Luapula provinces. Showing a persistence of hotspots in the south-west region over the years. Similarly, the spatial scan statistics exhibited a consistent location of the primary cluster of anemia in the south-west region, which could be attributed to poverty, malaria burden, dietary deficiencies and ITN gaps.

Regional disparities in anemia prevalence were observed in 2018 and 2024 ZDHS. Despite having a lower prevalence, 2024 exhibited higher prevalences at provincial and district levels, with a shift in high prevalence from Western to North-Western. Similarly, hotspot analysis suggested an increase in the hotspot area from those observed in Western to more pronounced ones extending towards North-Western and Luapula provinces. The emergence of North-Western and Luapula could be a reflection of persistent high malaria transmission intensity, coupled with limited access to health care and cross-border dynamics of the two provinces, which are known to influence anemia risk [36].

Contrary to the random distribution of anemia in the DRC [37], the study revealed a distinct spatial clustering in Zambia, suggesting anemia could be driven by localized environmental or regional factors such as malaria endemicity and regional food quality. Furthermore, similar significant clustering trends are observed in several SSA, including Ethiopia, Mali, Burkina Faso, and Nigeria [38–41].

Clearly, anemia in Zambia is a complex, multilevel problem. Despite individual biological determinants such as HIV and pregnancy remaining constant risks, the change in spatial hotspots and the effects of household and community-level factors suggest a national strategy may not be sufficient. Future interventions should be geographical targeted to address the socioeconomic barriers that prevent women from accessing both adequate nutrition and timely medical care.

## Conclusion

### Conclusion from findings

Anemia among women of reproductive age remains a persistent and significant moderate public health problem among WRA in Zambia. Through a multilevel approach, we demonstrated that anemia severity is influenced by complex interactions at the individual, household, and community levels. The factors identified in this study can be classified as risk factors of anemia which include, pregnancy, HIV status, financial barriers, and rural residency. Protective factors included being married. Thus, there is a need for integrated health services that link antenatal care, HIV management, and family planning with anemia screening and supplementation. The dynamic spatial distribution, that is, the movement of high prevalence clusters to North-Western and Luapula provinces imply drivers of anemia are evolving geographically. Signifying the need for geographically (subnation) targeted interventions and socio-economic sensitive policies which should include but not limited to active public health measurement coupled with nutritional education and interventions.

### Strengths and limitations of the study

The major strength of our study is in the use of a weighted, nationally representative sample from two ZDHS cycles, permitting a comprehensive comparison over the six years. Secondly, by employing a multilevel ordinal logistic regression, the study accounted for dependency in the data, providing reliable estimates and standard errors compared to the standard regression model. Furthermore, the integration of spatial analysis gives an understanding of the geographic variations of anemia in this population. Despite these strengths, the study has the following limitations. We are unable to establish temporal relationships due to the cross-sectional nature of the ZDHS data. The study only includes WRA who are alive at the time of the survey; deaths that may have been caused by anemia complications were not included, leading to survivor bias. Furthermore, important variables such as BMI, malaria infection, congenital infections, schistosomiasis, and/or hookworm are not available in DHS data [30, 31, 35, 42]. For privacy, the ZDHS masks subject location by displacing the GPS (latitude and longitude) data a total distance of 5 km for urban areas and 10 km for rural areas. This might bias our spatial distribution through underestimation of the hotspot intensity.

## Data Availability

The data underlying the results presented in the study are available from https://dhsprogram.com/

## Acknowledgments

I would like to thank Dr. Mable Mutengo for the support and encouragement in the entire research journey.

